# Ribavirin antiviral combination therapy in COVID 19, a single-center experience

**DOI:** 10.1101/2022.04.24.22274149

**Authors:** Budha O Singh, Gaurav Chikara, Prasan Kumar Panda, Yogesh Arvind Bahurupi, Sarama Saha, Venkatesh S. Pai

**Author notes:** **Corresponding author** Prasan Kumar Panda, Associate Professor, Dept. of Medicine, Sixth Floor, College Block, All India Institute of Medical Sciences (AIIMS), Rishikesh, India, 249203.

## Abstract

**Background:** COVID 19 infection has a similar clinical spectrum of disease presentation such as SARS and MERS in the past. These led to the assumption of the possibility to treat COVID 19 infection with antivirals which had been used to treat SARS and MERS.

**Methods:** A retrospective analysis was done on the data of SEV COVID Trial in symptomatic adult patients of COVID 19 infection with objectives to explore whether ribavirin antiviral combinations reduces the need of both noninvasive and invasive ventilators in treatment of COVID 19 infections.

**Results:** The patients were categorized as “Cohort A” consisting of 40 patients and “Cohort B” of 61 patients as Cohort A being the group of patients who received the standard therapy and Cohort B the group of patients who received the ribavirin combination therapy.

**Conclusion:** The study concluded that there was no statistically significant difference in regard to the need of noninvasive ventilation and invasive ventilation and also the development of multi-organ dysfunction in between the two Cohorts. Also, with progress of time, the proportion of patients with single organ dysfunctions in the two cohorts showed gradual recovery without any statistically significant differences.

## Introduction

COVID 19 infection caused by a novel beta-coronavirus, named Severe acute respiratory syndrome coronavirus 2 (SARS-CoV-2) has resulted into a pandemic and had led the WHO to declare it as a Global Sanitary Emergency on January 30, 2020 (1). The pathophysiologic process accompanying this highly transmissible infection primarily affects the respiratory system and may manifests as mild respiratory infection (may be responsible for 20-30% of the common colds), moderate, and finally to severe infections (2). However, it also has a wide clinical spectrum of presentation and can involve various organ and organ systems (3–6).

While the world has witnessed this pandemic in many waves, especially in India the 2nd wave has shown to have a sharp spike in and an exponentially increasing incidence of ICU admissions, organ impairments with the increased proportion of patients requiring high levels of O2 support and ventilator requirement in the recent times (7). As of now many antivirals have been proven to be of little or no benefits in the symptomatic recovery of the acute manifestations of COVID 19 infection.

Severe COVID 19 infections have also been attempted to treat with steroids (dexamethasone) and low molecular weight heparins and these have been proven to have some mortality benefit in moderate to severe cases of COVID 19 infections which also never led to a favorable outcome in mass scales before the progression of the severe pathophysiologic process (8). Several kinds of research have been done on the prospects of hydroxychloroquine, lopinavir/ritonavir, and ribavirin as individual agents for the treatment of COVID 19 infection. WHO Solidarity Trial Results has shown no benefit of these agents as indicated by no benefit on the overall mortality, initiation of ventilation, and duration of hospital stay (9).

This has led to the development of a retrospective cohort analysis of the COVID 19 patients who were enrolled in the SEV COVID trial (Safety and efficacy of antiviral combination therapy in symptomatic patients of COVID 19 infection – a randomized controlled trial) by Panda et al (Institutional Ethics Committee approval reference no. 218/IEC/IM/NF/2020) to analyze if the ribavirin combination therapy holds any benefit about reducing the need for ventilators and development of multi-organ dysfunctions in comparison to standard care..

## Material and Methods

The study was a retrospective record-based cohort study to find the effectiveness of antiviral combinations against the standard therapy from the data of the SEV COVID Trial which includes all adults who were hospitalized with COVID 19 infection. The study was approved by the Institutional Ethics Committee, AIIMS, Rishikesh (Letter No-AIIMS/IEC/21/526. Date: 09/10/2021). The population from the patients from this trial was segregated into the Cohort A which consists of patients who received the standard therapy (In SEV COVID Trial: Non-Severe A + Severe A) and Cohort B which consists of patients who received the ribavirin combination therapy (In SEV COVID Trial: Non-Severe B + Severe B + Severe C) after assessing individually for the fulfilment of the enrolment criteria (Table 1).

**Table 1:** Interventions received by the patients of the two cohorts.

| Table 1: Interventions received by the patients of the two cohorts |  |
| --- | --- |
| Cohort A: Standard therapies | Cohort B: Experimental therapies |
| 1. Strict Isolation<br>2. Standard Precautions (Hand hygiene, Cough Etiquette, Wear surgical mask)<br>3. Fluid Therapy<br>4. Supportive Pharmacotherapy (Antipyretic, Antiallergic, Cough Suppressant)<br>5. Oxygen supplementation (As required)<br>6. Invasive ventilation (As required) | 1. B1-(Hydroxychloroquine 400mg BD on day1 followed by 400 mg once daily + Ribavirin (2.4 gm orally as a loading dose followed by 1.2 gm orally every 12 hours) for 10 days + Standard Treatment)<br>2. B2- Lopinavir(400mg) + |
| 7. Antibiotic agents for other associated infections (according to 2019 ATS/IDSA guidelines for non-ICU and ICU patients) | Ritonavir (100mg) two tablets twice daily+ Ribavirin (2.4 gm orally as a loading dose followed by 1.2gm orally every 12 hours) for 10 days + Standard Treatment ) |
| 8. Vasopressor support |  |
| 9. Renal-replacement therapy |  |
| 10. Treatment of Comorbid Diseases |  |
| 11. Oseltamivir (75 mg BD) for patient who are tested positive for H1N1 |  |

Inclusion criteria were adults ≥ 18 years old who presented with signs and symptoms of COVID 19 infection and which was confirmed RT-PCR positive who were enrolled in the SEV COVID trial. Exclusion criteria were applied to patients who required non-invasive and invasive ventilation within 24 hours of the study, who were lost to follow-up during the trial period and who had refused consent for use of clinical data for research purposes.

The primary objectives set were to compare the need for ventilators (non-invasive and invasive and to compare the development of multi-organ dysfunction involving two or more organs in between interventional cohorts. The secondary objectives were to correlate the baseline patient characteristics with the above outcomes in between interventional cohorts, to determine and compare the average period from the date of enrolment to the need of ventilators (non-invasive and invasive), to determine and compare the average period from the date of enrolment to the development of acute organ dysfunction and to compare the need of invasive ventilators from non-invasive ones in between interventional cohorts.

All the baseline characteristics of selected patients were collected including demographics, symptoms, duration of illness, vitals at presentation and on follow-ups, pre-existing co-morbidities, baseline blood investigations and the parameters of specific organ dysfunction on presentation and on follow-ups, the requirement of invasive and noninvasive mechanical ventilation on a subsequent period of the study period. These were categorized based on the type of interventions the study groups have received.

Organ dysfunction of specific organ systems were defined as in respiratory System involvement by the requirement of oxygen support (radiological images not used since images were not uniformly available for all patients), hepatobiliary System by the elevation of serum SGOT of more than or equal to two times from baseline (chronic hepatitis ruled out from history and other routine investigations deemed necessary by the attending clinician), the involvement of renal system by elevation of serum creatinine of more than or equal to 1.5mg/dl after ruling out CKD from history (since baseline creatinine vales are not available more most patients) and elevated leukocyte count as a marker of sepsis after ruling out dehydration and other non-infective causes of leukocytosis. The outcome of the patient and time to symptomatic recovery in survivors (defined as the period from admission till resolution of symptoms or normalization of towards the baseline) were also noted.

Analysis were carried out such that categorical variables were presented in number and percentage (%), and continuous variables were presented as mean ± SD and median. Quantitative variables were compared using the Independent t-test (as the data sets were normally distributed) between two groups. Multivariate regression (logistic for categorical and linear for continuous dependent variables) was used to determine the significant predictor variables. A p-value of <0.05 was considered statistically significant.

## Results

From a sample of 111 patients, 101 was included and were distributed in Cohort A (40 patients) and Cohort B (61 patients) (Fig. 1). The patient characteristics in regard to age, co-morbidities, severity of disease at presentation, total leukocyte count, total and direct bilirubin, liver enzymes, urea and creatinine which were categorized as per the Cohorts were depicted in the table 2. There was no statistically significant difference in the proportion of individuals who develop the need for Non-Invasive Ventilation (NIV) and Invasive Ventilation (IV) in between the two Cohorts (Table 3). Similar observation was also made in regard to the secondary outcomes (Fig 2).

**Fig. 1:**
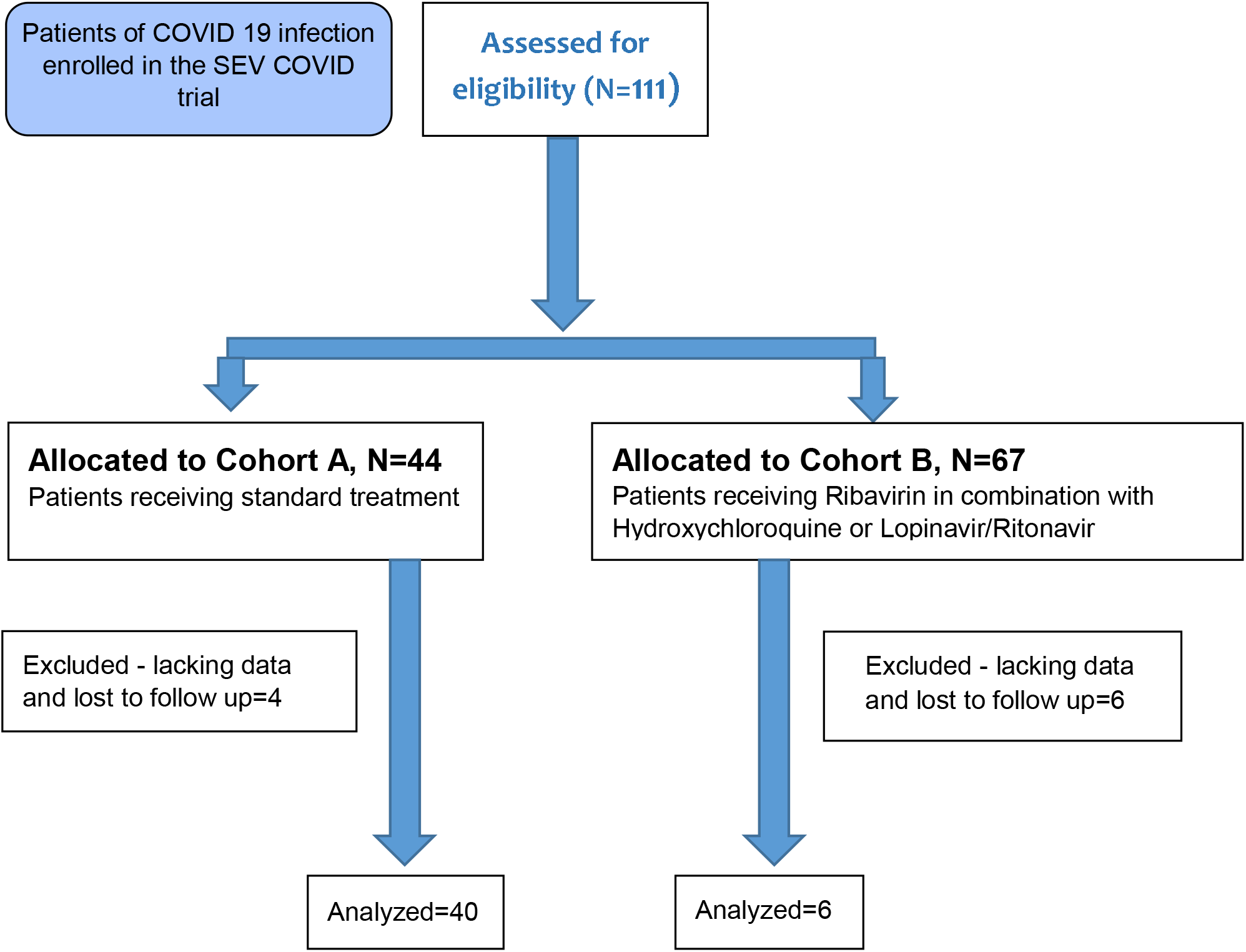
The study flow

**Table 2:**
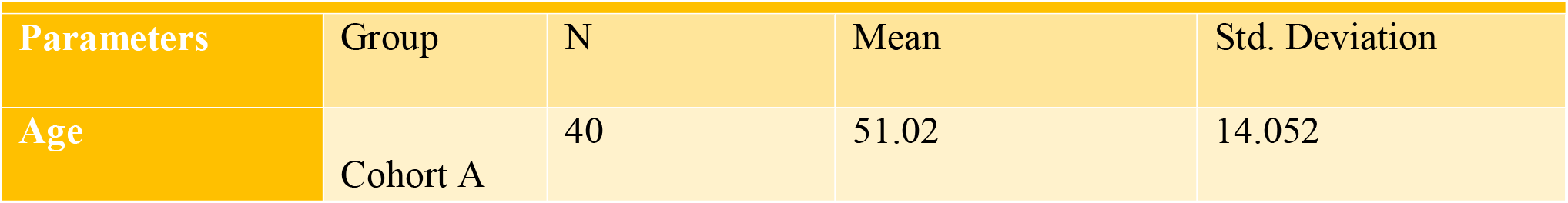
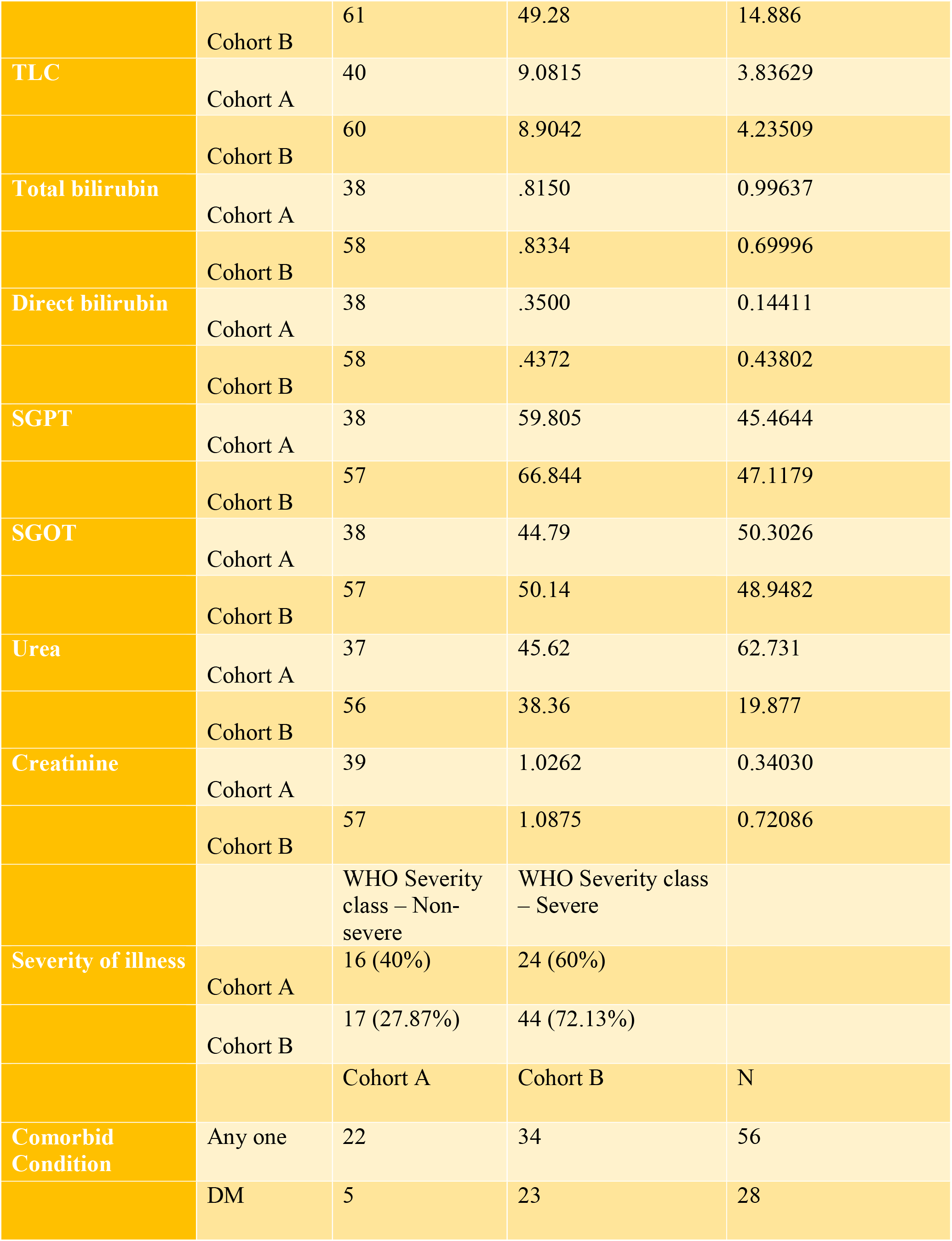
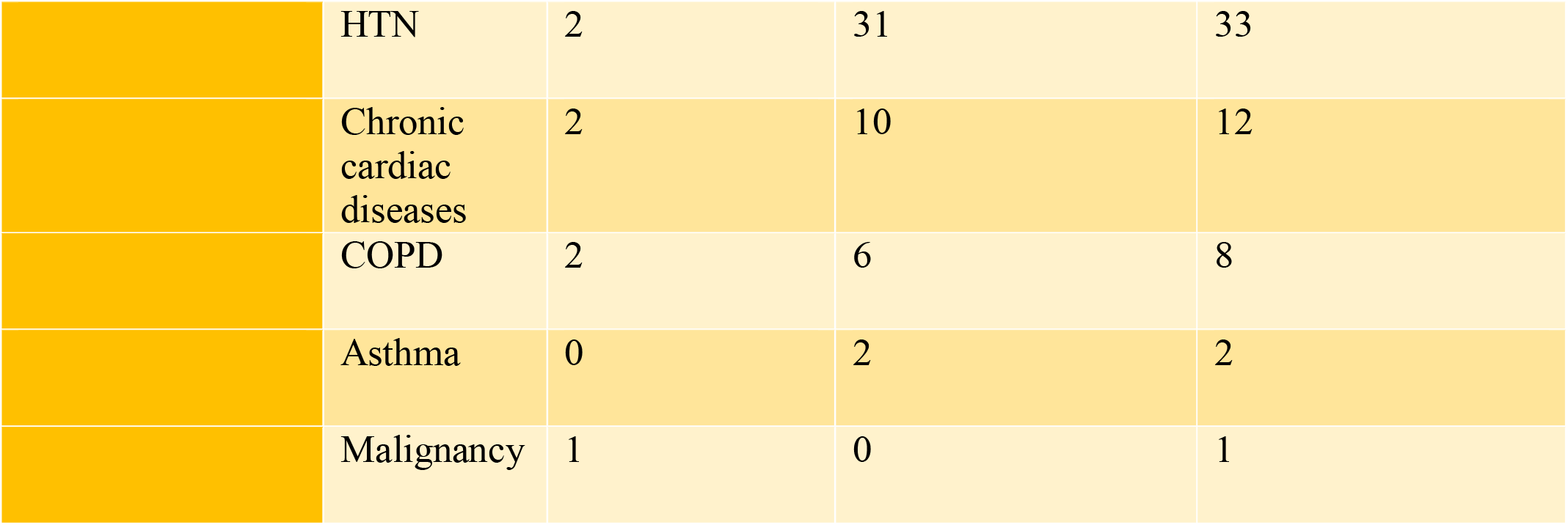
Demographic comparision of two cohorts.

**Table 3:** Non-Invasive Ventilation (NIV) and Invasive Ventilation (IV) requirement of Cohort A and B.

|  | Cohort A |  | Cohort B |  | P value |
| --- | --- | --- | --- | --- | --- |
|  | Count | Percentage | Count | Percentage |  |
| NIV | 5 | 12.5% | 11 | 18.03% | 0.456 |
| IV | 5 | 12.5% | 10 | 16.39% |  |
| Average Days in which requirement of NIV | 1.4 days |  | 2.54 days |  |  |
| Average Days in which requirement of IV | 3.2 days |  | 3.11 days |  |  |

**Fig 2:**
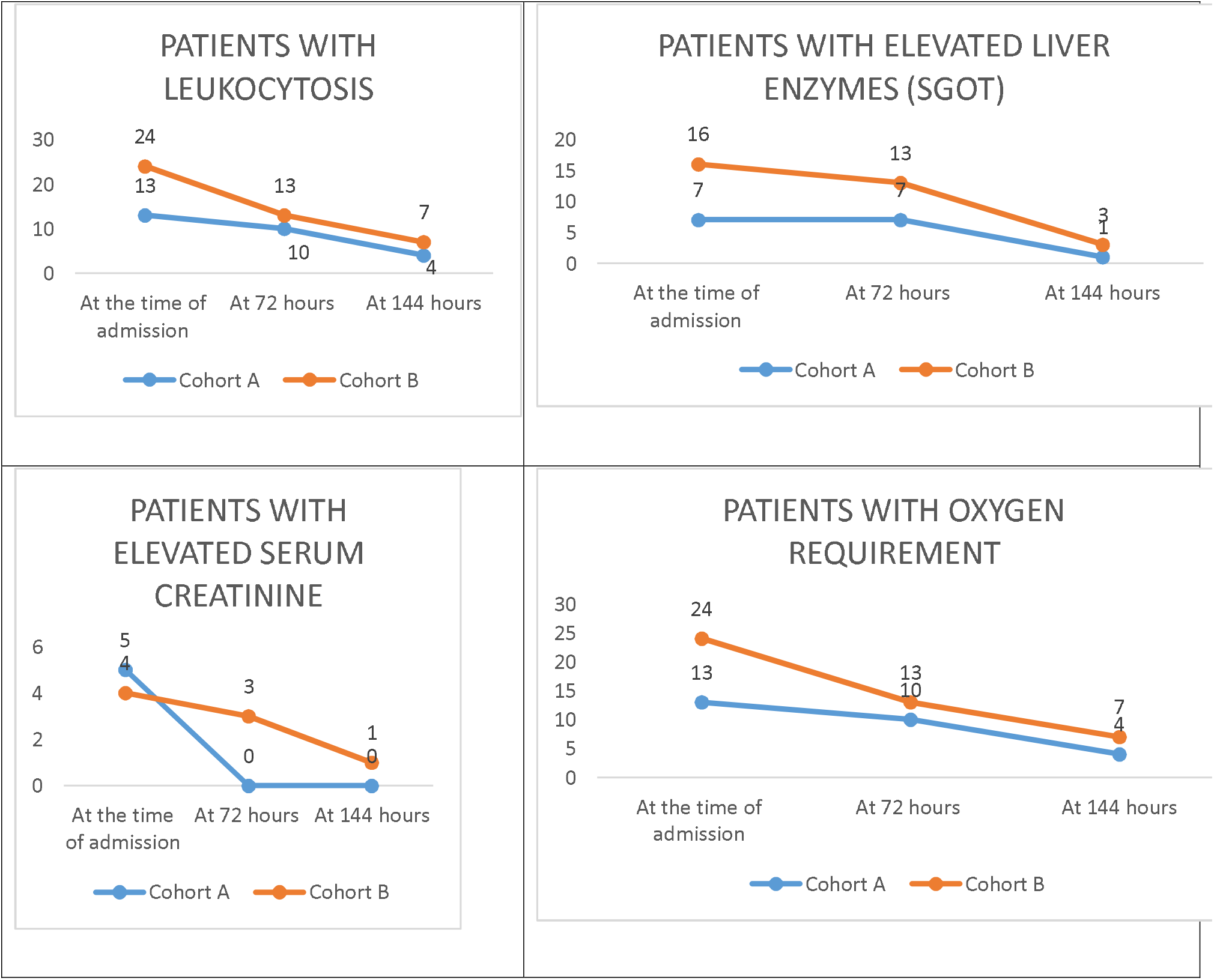
Time periods for development of acute organ dysfunctions between the two Cohorts

## Discussion

The study was done to assess whether ribavirin combination therapy poses any benefit in regard to the reduction of need of both NIV and IV requirement and the prevention/recovery from development of MODS as a result of COVID 19 infection. However, it could not establish the benefit of antiviral combinations over the supportive measure in respect to the outcomes examined. Type 2 diabetes mellitus, hypertension, chronic cardiac disease, COPD all in descending order of frequency were the major co morbid conditions with which the patients presented. This data is also supported by the previous findings from across the country as in the study by Ramanan Laxminarayan et al (10).

In regard to the need of NIV and IV between the two Cohorts, the study employed the previous recommendations concerning the use of this ventilator support. Studies like Nicolas Bonnet et al concluded a favorable outcome with use of HFNC to avoid the use of invasive ventilation (11). Whether its use might have resulted in a positive impact on the outcomes of present study is a question that was not answered by the parameters defined in this study. In regard to development of multi organ dysfunctions, the numbers in each sub-headings were too small for derivation of any statistically relevant and significant data.

The strict enrolment criteria of the primary study is a strong point in the times when studies on various antivirals and other therapeutic measures are taken on a large scale. While enrolling patients, the proportion of patients in both the cohorts were fairly comparable as it constitutes in the ratio of 2:3 between cohort A and B. The findings of our study were consistent with similar observations made in respect to epidemiological characteristics as in age groups, co-morbid conditions, and severity of illness.

Certain limitations may also be highlighted as the limited number of samples in the study which have impacted in deriving any statistically significant outcomes in the parameters used for the analysis. The data collected was dependent on the primary study and was not refined for a longer duration of observation due to various physical and logistic challenges imposed by the measures taken up during the pandemic. The results of the imaging studies such as X-ray and CT Thorax which is one of the primary modalities of classifying the severity of COVID 19 infection is avoided in the analysis due to lack of producible results and hence a possibility of bias is certainly present while classifying the severity of COVID 19 infection. And finally, to generalize the results of the effects of these drugs which were used in combination as a representative effect regarding the parameters of our study may also pose a certain bias.

In conclusion, there were no statistically significant difference in regard to the need of noninvasive ventilation (NIV) or invasive ventilation (IV) or development of multi-organ dysfunctions in between the two Cohorts by ribavirin combination therapy vs standard therapy.

## Data Availability

All data produced in the present study are available upon reasonable request to the authors

## Contributors

BOS contributed to the data collection, data analysis, and was involved in manuscript writing. GC, YAB, SS, and VSP contributed to the data collection and were involved in reviewing the draft. PKP gave the concept, interpreted analysis, critically reviewed the draft, and approved it for publication along with all authors.

## Data sharing

It will be made available to others as required upon requesting the corresponding author.

## Acknowledgment

None

